# Efficacy of Molnupiravir in reducing the risk of severe outcome in patients with SARS-CoV-2 infection: a real-life full-matched case-control study (SAVALO Study)

**DOI:** 10.1101/2024.09.09.24313305

**Authors:** Ivan Gentile, Riccardo Scotto, Maria Michela Scirocco, Francesco Di Brizzi, Federica Cuccurullo, Maria Silvitelli, Luigi Ametrano, Francesco Antimo Alfè, Daria Pietroluongo, Irene Irace, Mariarosaria Chiariello, Noemi De Felice, Simone Severino, Giulio Viceconte, Nicola Schiano Moriello, Alberto Enrico Maraolo, Antonio Riccardo Buonomo, Agnese Giaccone, Federico II COVID team

## Abstract

**Introduction:** Molnupiravir (MNP) is an orally administered prodrug which prevents disease progression in patients at high risk for severe COVID-19. We conducted a real-life case-control study on a cohort of outpatients, with Omicron SARS-CoV-2 infection to assess the effectiveness of MNP in reducing the occurrence of hospital admission, admission in intensive care unit (ICU) and death at day 28.

**Materials and methods:** Cases were enrolled among SARS-CoV-2 positive subjects that sought medical care during the first five days of symptoms from January 1st, 2022, to December 31st, 2022, and received MNP. Control participants were selected from a regional database among those who tested positive during the study period and did not receive antiviral treatment for SARS-CoV-2.

**Results:** 1382 patients were included (cases: 146, controls: 1236). Vaccinated patients showed lower risk of mortality and composite outcome (at least one among hospital admission, admission in ICU and all-cause death) compared to unvaccinated ones (0.6% vs 7.8%, p<0.001 and 2% vs 7.8%, p=0.001 respectively). After full-matching propensity score, MNP-treated subjects showed a lower incidence of composite outcome, while no effect was observed on the single outcomes. In the subgroup analysis according to the vaccination status, MNP proved effective in preventing all the outcomes among unvaccinated patients, while showed to reduce the risk of ICU admission both in vaccinated and unvaccinated patients.

**Conclusions:** Treatment with MNP proved effective in reducing the risk of composite outcome among outpatients with SARS-CoV-2 infection. The beneficial effect of MNP treatment in reducing progression is more pronounced in unvaccinated patients.

## Introduction

Since its emergence in late 2019, SARS-CoV-2 has continued to be a major cause of morbidity and mortality worldwide [1]. Several antiviral agents are currently available and are a cornerstone in the treatment of COVID-19, preventing the deterioration of patients, especially those at high risk for developing a severe disease. Although there are limited options for outpatient treatment, the introduction of effective oral antiviral therapies in the last two years has been a turning point for the early treatment of COVID-19 in ambulatory care settings [2, 3]. Molnupiravir (MNP; Lagevrio™, Merck & Co., Inc., Rahway, NJ, USA) is an orally administered ribonucleoside prodrug of N-hydroxycytidine (NHC) that has shown activity against influenza and other RNA viruses, including SARS-CoV-2. Once incorporated into viral RNA, it leads to an accumulation of deleterious errors, jeopardizing the replication capacity of the virus [4, 5]. Supported by data from the phase 3, double-blind, randomized, placebo-controlled MOVe-OUT trial, which showed a risk of hospitalization or death reduced by 30% in at-risk outpatients treated with MNP compared to placebo, MNP received emergency use authorization (EUA) by the U.S. Food and Drug Administration (FDA) in December 2021 for the treatment of non-hospitalized patients with symptomatic COVID-19 at high risk of progression to severe COVID-19 [4, 6]. However, the MOVe-OUT trial was conducted among unvaccinated patients before the emergence of the Omicron (B.1.1.529) variant and subsequent sub-lineages. Therefore, collecting real-life data on the effectiveness of oral antiviral agents in a mostly immunized population with predominant Omicron variants infections has become a research priority.

In light of this, the present case-control study aims to assess the e^icacy of MNP in preventing hospital admission, admission to intensive care unit (ICU) and death in a real-life population, mostly vaccinated or possessing hybrid immunity, with SARS-CoV-2 Omicron variants infection.

## Materials and methods

### Study design and population

This study was designed as a retrospective case-control analysis. Participants were required to meet the following inclusion criteria: a positive SARS-CoV-2 test between January 2022 and December 2022, an age of 18 years or older, and no prior treatment with COVID-19 therapies such as nirmatrelvir/ritonavir, remdesivir, sotrovimab, tixagevimab/cilgavimab, or other anti-SARS-CoV-2 monoclonal antibodies, either alone or in combination with MNP.

Cases in this study were defined as consecutive individuals who sought outpatient care at the Infectious Diseases Unit of Federico II University in Naples, Italy, and were treated with molnupiravir within five days of symptom onset. All patients who received treatment had risk factors for developing severe COVID-19, as outlined by the Italian Medicines Agency (AIFA). These risk factors included being over 65 years of age, having solid or haematological cancer, immunodeficiency, chronic liver or kidney disease, cardiovascular disease, uncontrolled diabetes mellitus, hemoglobinopathies, neurological disorders, chronic obstructive pulmonary disease (COPD), or severe obesity (BMI ≥ 30) [7]

Control participants were selected from a regional database of individuals who tested positive for SARS-CoV-2 during the study period. Those who had received any specific treatment for COVID-19 were excluded. Controls were identified and validated through telephone interviews designed by the authors and conducted by healthcare professionals (see Supplementary File 1 for the full interview script).

### Data collection and definitions

We collected comprehensive data on patients’ demographic characteristics (age, sex), comorbid conditions (such as diabetes mellitus, hypertension, heart disease, lung disease, immunodeficiency, kidney or hepatic disorders, neurological conditions, and obesity), vaccination status, and the predominant SARS-CoV-2 variant in our region during each phase of the study period. Patients were classified as fully vaccinated if they had received at least two doses of a COVID-19 vaccine. For each patient, we calculated the Monoclonal Antibody Screening Score (MASS), originally developed to rapidly identify and stratify patients according to their risk of hospitalization, thereby determining their priority for monoclonal antibody treatment [8]. Additionally, we formulated a simplified Comorbidity Score: patients with three or more comorbidities were assigned 2 points, those with one or two comorbidities received 1 point, and those without any comorbidities were assigned 0 points.

### Propensity score matching

We performed a propensity score (PS)-matched analysis to mitigate selection bias and ensure an equitable distribution of confounding variables between cases and controls [9]. Our approach to PS methods followed the guidelines recommended by Eikenboom et al., aimed at enhancing the quality of research on the effectiveness of antimicrobial therapies [10].

A PS model was developed for each subject, assigning a probability of receiving molnupiravir based on baseline characteristics such as age, sex, vaccination status, comorbid conditions, MASS, simplified Comorbidity Score, and the predominant SARS-CoV-2 variant. Probabilities were calculated using probit regression. The balance of PS matching was evaluated by measuring the absolute standardized mean difference (SMD) of covariates, both continuous and categorical, between the groups. A value below 0.1 was deemed to indicate an acceptable balance.

We assessed various matching algorithms, including greedy matching (with different ratios and calipers), optimal matching, and full matching [11]. The latter yielded the optimal balance while preserving the entire sample size. This approach allocated each unit in the sample to a subclass, with subclasses containing either one treated unit and one or more control units or one control unit and one or more treated units. The number of subclasses and the allocation of units were optimized to minimize the sum of absolute within-subclass distances in the matched sample [12].

### Outcomes

The primary outcomes of the study were the proportions of hospital admissions, intensive care unit (ICU) admissions, and 28-day all-cause mortality among both the treated individuals and the propensity-matched untreated individuals. Additionally, we evaluated a composite outcome, defined as the occurrence of at least one of the above-mentioned events. The follow-up period extended to 28 days from the onset of symptoms, or the first positive SARS-CoV-2 test.

### Statistical analysis

In this study, categorical variables are presented as counts and percentages, while continuous variables are reported as medians with interquartile ranges (IQRs). The Pearson’s χ2 test was employed to assess differences in baseline characteristics between groups for categorical variables, and the Mann-Whitney U test was used for continuous variables.

After matching, the effect of molnupiravir (MNP) on the outcomes of interest was estimated. The chosen estimand was the average treatment effect on the treated (ATT), which represents the average effect of treatment among those who received it. This estimand addresses the following fundamental question: should medical providers refrain from withholding treatment from those currently receiving it? [13]. We estimated marginal effects, comparing the expected potential outcomes under treatment to those under control, and expressed these as odds ratios (ORs) with 95% confidence intervals (CIs). Estimation was conducted using g-computation with a cluster-robust standard error to account for pair membership, focusing on binary outcomes and incorporating covariates via logistic regression [14]. Including covariates in the outcome model after matching serves several purposes: it can enhance precision in the effect estimate, reduce bias due to residual imbalance, and provide a “doubly robust” effect estimate. This means the estimate remains consistent if either the matching sufficiently reduces covariate imbalance or if the outcome model is correctly specified [15].

All analyses were performed using R, version 4.1.0 (R Core Team), with the following packages:

*MatchIt, cobalt*, and *marginal effects*.

### Secondary analysis

A moderation analysis was performed to assess whether the treatment effect differed across varying levels of another variable, specifically vaccination status. This was achieved by carrying out matching on the full dataset to ensure balance within each subgroup of the moderating variable [16].

### Ethical approval and consent to participate

This study received approval from the ethics committee “Comitato Etico Università Federico II-A.O.R.N. A. Cardarelli” (protocol number 0015191, 22nd March 2023). Informed consent was obtained from all patients included in the study. For control patients, consent was obtained via telephone interview, a method that was rigorously reviewed and approved by the ethics committee to ensure it met all ethical standards. Informed consent from case patients was obtained in written form. Records of the telephone interviews with control participants, including their verbal consent, are securely stored by the study’s data manager at the Department of Clinical Medicine and Surgery, University of Naples Federico II, Via Sergio Pansini 5, 80131, Naples, Italy. These records are available for review by any relevant authority upon reasonable request.

## Results

The study included a total of 1382 patients who met the inclusion/exclusion criteria. This comprised 146 cases (10.6%) and 1236 controls (89.4%). The baseline characteristics of these cases and controls are detailed in *Table 1*. Patients treated with molnupiravir exhibited a more complex clinical profile. Specifically, they were older (p<0.001), they more frequently had hypertension, chronic heart disease, chronic kidney disease (p<0.001), COPD (p=0.024) and more often had immunodeficiency (p<0.001). These patients also exhibited a higher comorbidity score (p<0.001) and a higher MASS score (p<0.001).

**Table 1.** Baseline features of pre-matched sample (N=1382).

|  | **Molnupiravir** | **No therapy** | **p-value** |
| --- | --- | --- | --- |
| N | 146 | 1236 |  |
| Male Sex (n, %) | 77 (52.7) | 551 (44.6) | 0.074 |
| Age (median [IQR]) | 70.00 [59.00, 79.00] | 57.00 [46.75, 68.00] | <0.001 |
| Vaccination (%) | 138 (94.5) | 1142 (92.4) | 0.446 |
| Diabetes (%) | 13 (8.9) | 115 (9.3) | 0.995 |
| Hypertension (%) | 52 (35.6) | 245 (20.6) | <0.001 |
| Chronic heart disease (%) | 60 (41.1) | 200 (16.2) | <0.001 |
| COPD (%) | 18 (12.3) | 84 (6.8) | 0.024 |
| CKD (%) | 11 (7.5) | 24 (1.9) | <0.001 |
| Obesity (%) | 23 (15.8) | 145 (11.7) | 0.203 |
| Liver disease (%) | 4 (2.7) | 14 (1.1) | 0.217 |
| Neurological disease (%) | 6 (4.1) | 46 (3.7) | 0.998 |
| Immunodeficiency (%) | 71 (48.6) | 80 (6.5) | <0.001 |
| Comorbidity Score (median [IQR]) | 1.00 [1.00, 1.00] | 0.00 [0.00, 1.00] | <0.001 |
| MASS score (median [IQR]) | 5.00 [3.00, 6.75] | 1.00 [0.00, 3.00] | <0.001 |
| Predominant variant (%) |  |  | <0.001 |
| - omicron (not specified) | 0 (0.0) | 123 (10.0) |  |
| - omicron_BA.1 | 18 (12.3) | 212 (17.2) |  |
| - omicron_BA.2 | 128 (87.7) | 271 (21.9) |  |
| - omicron_BA.5 | 0 (0.0) | 627 (50.7) |  |
| - omicron_BQ.1 | 0 (0.0) | 3 (0.2) |  |

*IQR: interquartile range; COPD: chronic obstructive pulmonary disease; CKD: chronic kidney disease*

In total, 23 patients (1.7%) required hospitalization, and 1 patient (0.1%) was admitted to ICU. Only 1 out of 146 (0.7%) patients treated with MNP died, while 15 (1.2%) patients among controls died. However, no significant differences were observed in any of the outcomes under evaluation between the treated and untreated groups (*Table 2*).

**Table 2.** Outcome rates among cases and controls.

|  | **Molnupiravir** | **No therapy** | **p-value** |
| --- | --- | --- | --- |
| n | 146 | 1236 |  |
| Hospital admission (%) | 3 (2.1) | 20 (1.6) | 0.962 |
| ICU admission (%) | 0 (0.0) | 1 (0.1) | 1.000 |
| Death (%) | 1 (0.7) | 15 (1.2) | 0.876 |
| Composite outcome (%) | 3 (2.1) | 31 (2.5) | 0.959 |

However, after full-matching propensity score (see Supplementary Figure 1 and Supplementary Figure 2 for covariate balance and distribution of PS between groups), MNP showed to be effective in reducing the rate of the composite outcome, while no effect on hospital admission or death alone was observed (*Table 3*).

**Table 3.** Effect of MNP on different outcomes after full-matched propensity score.

|  | Point estimate (OR) | Standard Error | 95% Confidence Interval | P value |
| --- | --- | --- | --- | --- |
| Composite outcome | 0.353 | 6.2 | 0.155-0.805 | 0.013 |
| Hospital admission | 0.744 | 0.4 | 0.132-4.18 | 0.737 |
| ICU admission | * | * | * | * |
| Death | 0.958 | 0.3 | 0.701-1.31 | 0.787 |

\*Algorithm did not converge (too few events).

When comparing outcomes after stratification for vaccination status, we observed that patients who has received the SARS-CoV-2 vaccine had lower death rate compared to unvaccinated patients (p<0.001) and a significantly reduced risk of the composite outcome (p=0.001) (*Table 4*).

**Table 4.**
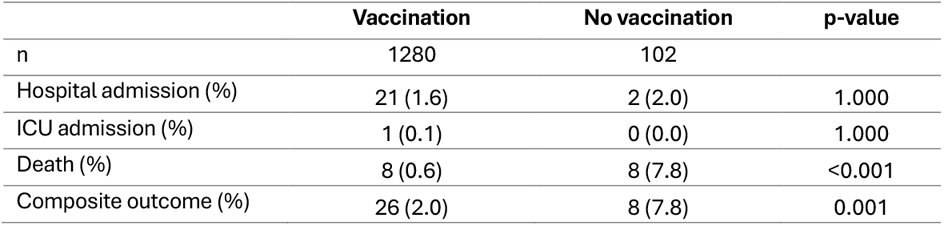
Outcome rates among included patients according to vaccination status.

|  | Vaccination | No vaccination | p-value |
| --- | --- | --- | --- |
| n | 1280 | 102 |  |
| Hospital admission (%) | 21 (1.6) | 2 (2.0) | 1.000 |
| ICU admission (%) | 1 (0.1) | 0 (0.0) | 1.000 |
| Death (%) | 8 (0.6) | 8 (7.8) | <0.001 |
| Composite outcome (%) | 26 (2.0) | 8 (7.8) | 0.001 |

When stratifying treated and untreated patients according to their vaccination status, none of the evaluated outcomes showed a significative reduction in the MNP arm (see Supplementary Table 1). In the subgroup analysis conducted among both cases and controls who received the SARS-CoV-2 vaccine, MNP showed to reduce the risk of ICU admission both in vaccinated and unvaccinated subjects after full-matched propensity score analysis (p<0.001) (*Table 5*). Notably, MNP was also found to be effective in reducing the risk of hospital admission, death and composite outcome (p<0.001) in non-vaccinated individuals.

**Table 5.**
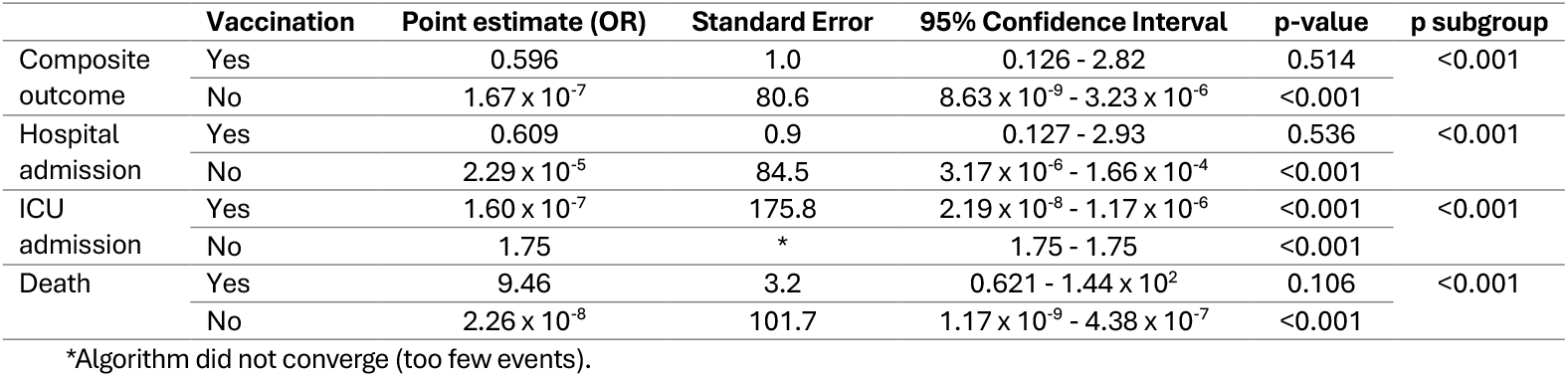
Effect of MNP on different outcomes after full-matched propensity score, according to vaccination status.

|  | Vaccination | Point estimate (OR) | Standard Error | 95% Confidence Interval | p-value | p subgroup |
| --- | --- | --- | --- | --- | --- | --- |
| Composite outcome | Yes | 0.596 | 1.0 | 0.126 - 2.82 | 0.514 | <0.001 |
| | No | $1.67 \times 10^{-7}$ | 80.6 | $8.63 \times 10^{-9}$ - $3.23 \times 10^{-6}$ | <0.001 | |
| Hospital admission | Yes | 0.609 | 0.9 | 0.127 - 2.93 | 0.536 | <0.001 |
| | No | $2.29 \times 10^{-5}$ | 84.5 | $3.17 \times 10^{-6}$ - $1.66 \times 10^{-4}$ | <0.001 | |
| ICU admission | Yes | $1.60 \times 10^{-7}$ | 175.8 | $2.19 \times 10^{-8}$ - $1.17 \times 10^{-6}$ | <0.001 | <0.001 |
|  | No | 1.75 | * | 1.75 - 1.75 | <0.001 |  |
| Death | Yes | 9.46 | 3.2 | 0.621 - $1.44 \times 10^2$ | 0.106 | <0.001 |
| | No | $2.26 \times 10^{-8}$ | 101.7 | $1.17 \times 10^{-9}$ - $4.38 \times 10^{-7}$ | <0.001 | |

\*Algorithm did not converge (too few events).

## Discussion

In this retrospective, PS-matched cohort study, we evaluated the real-life effectiveness of molnupiravir in preventing hospitalization, ICU admission and death among outpatients with Omicron SARS-CoV-2 infection in a highly vaccinated population. The full-matching propensity score provided a fair and balanced comparison between cases and controls. We collected data on the outcomes of MNP-treated patients and untreated patients during the same period to achieve a relative homogeneity in the infecting variants of SARS-CoV-2 between the two groups. Specifically, Omicron BA.1, BA.2 and BA.5 were the most prevalent variants: among cases, we observed a polarization around BA.2 (87.7%), while the variants were more evenly distributed among controls with a prevalence of BA.5 (50.7%).

The vast majority of both cases and controls in our study (>90%) had received at least two doses of SARS-CoV-2 vaccine. Given the high rate of immunization of our cohort and the prevalence of Omicron variants, whose virulence is well-known to be inferior to previous SARS-CoV-2 variants, a low rate of disease progression and death was expected, in line with worldwide epidemiological data [17]. Notably, although not statistically significant, 15 deaths were reported among the controls, while only one death was recorded in the MNP-receiving group. This is a remarkable finding considering that the latter group included patients with a frailer clinical profile, significantly older mean age and higher comorbidity scores, as reported in *Table 1*.

A more favorable disease course was confirmed after PS analysis, with a lower incidence of the composite outcome (at least one among hospital admission, ICU admission and death) in the MNP-treated group (p=0.013); however, no significant reduction in separate rate of hospital admission or death was observed. Our results are partially consistent with those of other cohorts.

Actually, the large, open-label, randomized controlled PANORAMIC trial assessed the efficacy of early treatment with molnupiravir versus placebo among a highly vaccinated population (94% of participants had received at least three doses of a SARS-CoV-2 vaccine). The trial demonstrated earlier symptoms alleviation and faster viral load decline in the MNP-receiving group but no significant reduction in hospital admission and mortality rate (which was very low, around 1% in both treatment arms) [18]. We reported a similar result in our study after PS-matching, with no significant difference in hospitalization and mortality rate, although we found MNP to be effective in reducing the occurrence of composite outcome in our highly vaccinated cohort.

Moreover, Wong et al., in a retrospective, propensity-score matched analysis on a cohort of hospitalised patients with COVID-19 not requiring oxygen therapy on admission, found that early administration of MNP significantly reduce the risk of all-cause mortality and disease progression

(hazard ratio, HR 0.60, 95% CI 0.52-0.69) [3]. Similarly, a propensity score-matched retrospective study conducted among U.S. veterans showed a lower combined 30-day risk of hospitalization or death in molnupiravir-treated participants aged ≥ 65 years versus no treatment (relative risk, RR 0.67, 95% CI 0.46-0.99) [19].

Conversely, some studies reported no significant effect of MNP, as shown by Yip et al. in their retrospective propensity-score matched analysis [20]. Accordingly, Najjar-Debbiny et al. reported a nonsignificant reduced risk of the composite outcome (HR 0.83, 95% CI 0.57-1.21) in the MNP-treated group. However, subgroup analyses showed that MNP was associated with a significant decrease in the risk of the composite outcome in older patients, females, and patients with inadequate COVID-19 vaccination [21]. A newly published systematic review of nine real-world studies reported that MNP was effective in reducing the risk of severe COVID-19 outcomes, particularly among older age groups. The authors reported as potential bias a difference in the baseline characteristics of the MNP-treated group versus the untreated group, with the former including generally older participants and with more comorbidities than controls. Therefore, they inferred that the actual effectiveness of molnupiravir may have been underestimated [22]. We might draw the same inference for our cohort, where the cases were consistently older and more vulnerable than controls, even after minimizing the differences in the baseline features between the two groups by PS-matching. Additionally, all the studies included in the above-mentioned systematic review were conducted when the Omicron subvariants were predominant and a relevant part of the population already had prior immunity to SARS-CoV-2 [22]. It is worth mentioning that, although the MOVe-OUT trial only included unvaccinated patients, in the subgroup analysis of patients with evidence of previous SARS-CoV-2 infection, molnupiravir showed no better outcome compared to placebo [4]. Given the high rate of vaccination against COVID-19 among the general population, real-world studies play a crucial role in evaluating the real advantage of MNP employment in patients with prior SARS-CoV-2 immunity (either acquired through vaccination or past infection).

In our study, we observed a significantly higher incidence of death (7.8% vs 0.6%, p<0.001) and composite outcome (7.8% vs 2%, p=0.001) among unvaccinated patients, than in vaccinated patients (*Table 4*). This finding supports the evidence that vaccination against SARS-CoV-2 is highly effective in preventing clinical deterioration and death regardless of the immune escape of new variants and their limited virulence compared to the wild-type virus.

As expected, the most profound effect was observed among unvaccinated individuals, for whom molnupiravir proved effective in reducing the occurrence all evaluated outcomes, with a remarkably higher benefit from the antiviral therapy compared to fully vaccinated patients (see *Table 5*). Among the latter, MNP reduced the risk of the composite outcome and of hospitalization, but not in a statistically significant fashion. Nonetheless, MNP retained significant efficacy in preventing ICU admission among fully vaccinated subjects. This underscores a potential benefit of MNP regardless of vaccination status. As is the case with many medical interventions, there is likely to be a gradient of benefit for treatment with MNP, with the greatest impact shown in the subjects at highest risk for progression and without prior immunization [23].

The apparent lack of effectiveness reported by some studies might partly be related to its availability in earlier days when prescribing criteria were more stringent, and molnupiravir was preferentially assigned to more frail and polymedicated patients than those who received nirmatrelvir/ritonavir, perhaps because of the multiple drug-drug interactions associated with the latter [20].

Essentially, our results, along with recent real-life literature, suggest that MNP may be effective in reducing the risk of unfavorable outcomes in patients with SARS-CoV-2 infection. Unfortunately, after the completion of our study the Scientific Technical Commission of the Italian Drug Agency (AIFA) decided to suspend the use of molnupiravir in March 2023, following the negative opinion issued by the European Medicines Agency (EMA) [24, 25]. This decision leaves European patients who need oral antivirals but cannot take nirmatrelvir/ritonavir (e.g., due to insurmountable drug-to-drug interactions) with no other viable therapeutic options apart from a short course of remdesivir, which, however, incurs hospitalization costs. According to the CDC, molnupiravir remains a second-line treatment option for patients with SARS-CoV-2 infection at risk for severe disease who cannot be treated with nirmatrelvir/ritonavir or remdesivir, and MNP continues to be administered in the United States [26]. Additionally, the WHO’s living guidelines on COVID-19 conditionally recommend the use of MNP for patients with non-severe COVID-19 at the highest risk of hospitalization (excluding pregnant and breastfeeding women, and children) [27]

Major strengths of our study are the large number of controls and the use of PS full-matching, which allows to estimate a treatment effect which is ideally free of confounding due to the measured covariates. Indeed, this algorithm ensures a more balanced matching given that all individuals are retained, minimizing the confounding effect of a marked disproportion between the two groups in a key variable at baseline, namely the immunodeficiency status that is present in 48.6% vs 6.5% among cases and controls, respectively. Especially regarding the impact of MNP on vaccinated and unvaccinated patients, the PS analysis enabled to detect a substantial improvement of all the evaluated outcomes which had not previously emerged in the baseline statistical analysis. Moreover, our study can contribute to real-world evidence in a context wherein it has become more difficult to set up clinical trials that can keep up with an evolving viral landscape of variants. The final message is in line with the updated living guidelines by the World Health Organization– November 2023 – recommending MNP for infected patients at high risk of progression.

We recognize that our study has nonetheless some limitations. Firstly, our study is subject to all the biases inherent to its retrospective study design. While data for the cases were collected prospectively, data for the controls, who represent a large percentage of all the included patients (89.4%), were retrieved retrospectively. Since controls were identified through telephone interviews directly or via their relatives, some may not have accurately reported baseline features (e.g., vaccination status) or outcomes. For instance, hospital admissions beyond the predefined follow-up window might have been reported as having occurred earlier due to recall bias. Despite efforts to track relatives of deceased patients, it is possible that a significant percentage of controls (or their relatives) who had an unfavourable outcome declined to be interviewed. However, given the large number of controls included in the study, this bias may have been mitigated.

## Conclusions

Our real-life study demonstrated that, after full-matching propensity-score, molnupiravir can reduce the risk of composite outcome (at least one among hospitalisation, ICU admission and all-cause death) at day 28, irrespective of vaccination status, in a highly vaccinated outpatients infected with SARS-CoV-2 Omicron variants.

The beneficial effect of MNP treatment in reducing progression is more pronounced in unvaccinated patients. Its real effectiveness, however, might have been underestimated due to residual unmeasured confounding that was not adjusted by PS

## Data Availability

The datasets generated and/or analysed during the current study, along with the studys Case Report Forms (CRFs), the written informed consent from case participants, and the records of the telephonic interviews conducted with control participants, including their verbal informed consent, are stored by the studys data manager at the Department of Clinical Medicine and Surgery, University of Naples Federico II, Via Sergio Pansini 5, 80131, Naples, Italy. These can be retrieved and reviewed upon request by any relevant authority.

## Ackowledgements

Federico II COVID Team: Francesco Antimo Alfè^1^, Luigi Ametrano^1^, Anna Borrelli^2^, Antonio Riccardo Buonomo^1^, Ferdinando Calabria^1^, Giuseppe Castaldo^3^, Letizia Cattaneo^1^, Maria Rosaria Chiariello^1^, Mariarosaria Cotugno^1^, Federica Cuccurullo^1^, Alessia d’Agostino^1^, Noemi De Felice^1^, Dario Diana^1^, Francesco Di Brizzi^1^, Giovanni Di Filippo^1^, Isabella Di Filippo^1^, Antonio Di Fusco^1^, Federico Di Panni^1^, Gaia Di Troia^1^, Nunzia Esposito^1^, Mariarosaria Faiella^1^, Lidia Festa^1^, Maria Foggia^1^, Maria Elisabetta Forte^1^, Ludovica Fusco^1^, Antonella Gallicchio, Gianpaolo Gargiulo^4^, Ivan Gentile^1^, Antonia Gesmundo^1^, Agnese Giaccone^5^, Carmela Iervolino^1^, Irene Irace^1^, Antonio Iuliano^1^, Federica Licciardi^1^, Giuseppe Longo^2^, Matteo Lorito^6^, Alberto Enrico Maraolo^1^, Simona Mercinelli^1^, Fulvio Minervini^1^, Giuseppina Muto^1^, Mariano Nobile^1^, Daria Pietroluongo^1^, Biagio Pinchera^1^, Giuseppe Portella^7^, Laura Reynaud^1^, Alessia Sardanelli^1^, Marina Sarno^1^, Simone Severino^1^, Maria Silvitelli^1^, Nicola Schiano Moriello^1^, Maria Michela Scirocco^1^, Fabrizio Scordino^1^, Riccardo Scotto^1^, Stefano Mario Susini^1^, Anastasia Tanzillo^1^, Grazia Tosone^1^, Emilia Trucillo^1^, Ilaria Vecchietti^1^, Giulio Viceconte^1^, Emanuela Zappulo^1^, Giulia Zumbo^1^

^1^Department of Clinical Medicine and Surgery – Section of Infectious Diseases, University of Naples Federico II, Naples, Italy

^2^Hospital Direction, University of Naples Federico II, Naples, Italy

^3^Department of Medical Biotechnology and Molecular Medicine, University of Naples Federico II, Naples, Italy

^4^Department of Clinical Medicine and Surgery, University of Naples Federico II, Naples, Italy

^5^AORN Ospedali dei Colli, Cotugno Hospital, Department of Infectious Diseases, Unit of Geriatric Infectious Diseases, Naples, Italy

^6^University of Naples Federico II, Naples, Italy

^7^Department of Translational Medical Sciences, University of Naples Federico II, Naples, Italy

## Availability of data and materials

The datasets generated and/or analysed during the current study, along with the study’s Case Report Forms (CRFs), the written informed consent from case participants, and the records of the telephonic interviews conducted with control participants, including their verbal informed consent, are stored by the study’s data manager at the Department of Clinical Medicine and Surgery, University of Naples Federico II, Via Sergio Pansini 5, 80131, Naples, Italy. These can be retrieved and reviewed upon request by any relevant authority.

## Competing interests

Prof. IVAN GENTILE reports personal fees from MSD, AbbVie, Gilead, Pfizer, GSK, SOBI, Nordic/Infecto Pharm, Angelini and Abbott, as well as departmental grants from Gilead and support for attending a meeting from Janssen, outside the submitted work.

All other authors have no competing interests to declare

## Funding

This study was funded by the Campania Region as part of the European Fund for Regional Development (Fondo Europeo Sviluppo Regionale – FESR) for 2014-2020, under the project titled ‘Impact of Early Anti-SARS-CoV-2 Treatment in Vaccinated Subjects on Clinical Progression and Onset of Long-COVID (SAVALO Study)’. The funds received have been utilized to engage nursing staff who were responsible for conducting telephone interviews with control participants who opted to participate, and for collecting data on paper-based Case Report Forms (CRFs). The funds will also be allocated for publication charges, if necessary.

## Declaration of generative AI and AI-assisted technologies in the writing process

During the preparation of this work the authors used Microsoft© Copilot, powered by Chat-GPT IV, after the completion of the first draft, in order to improve readability and the English grammar of the paper. After using this tool, the authors reviewed and edited the content as needed and take full responsibility for the content of the published article

## Notes

### Author Declarations

This study received approval from the ethics committee "Comitato Etico Università Federico II-A.O.R.N. A. Cardarelli" (protocol number 0015191, 22nd March 2023).

