## Supplementary figures and images for "Efficacy of Molnupiravir in reducing the risk of severe outcome in patients with SARS-CoV-2 infection: a real-life full-matched case-control study (SAVALO Study)"

### Supplementary figure 1

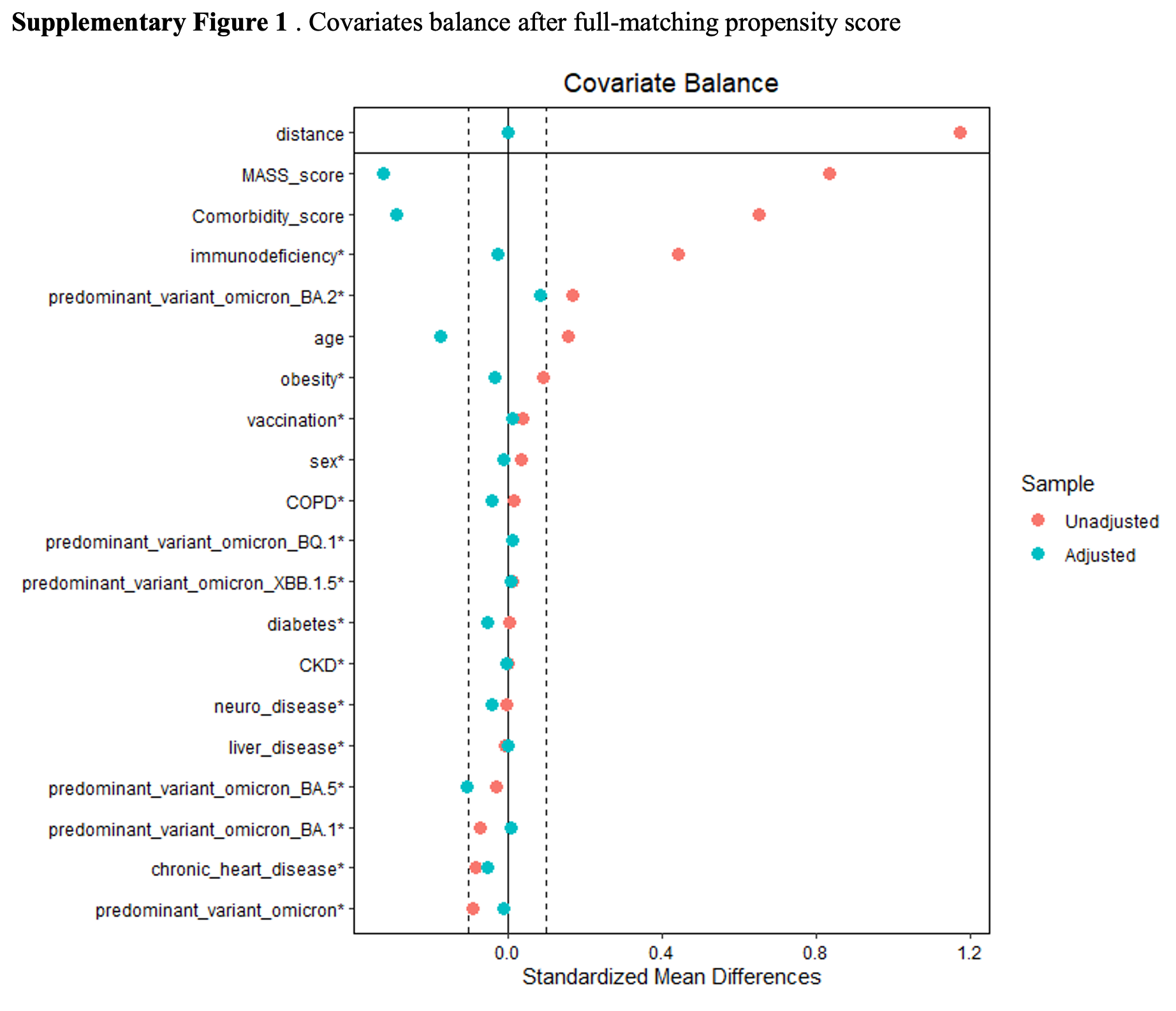

### Supplementary figure 2

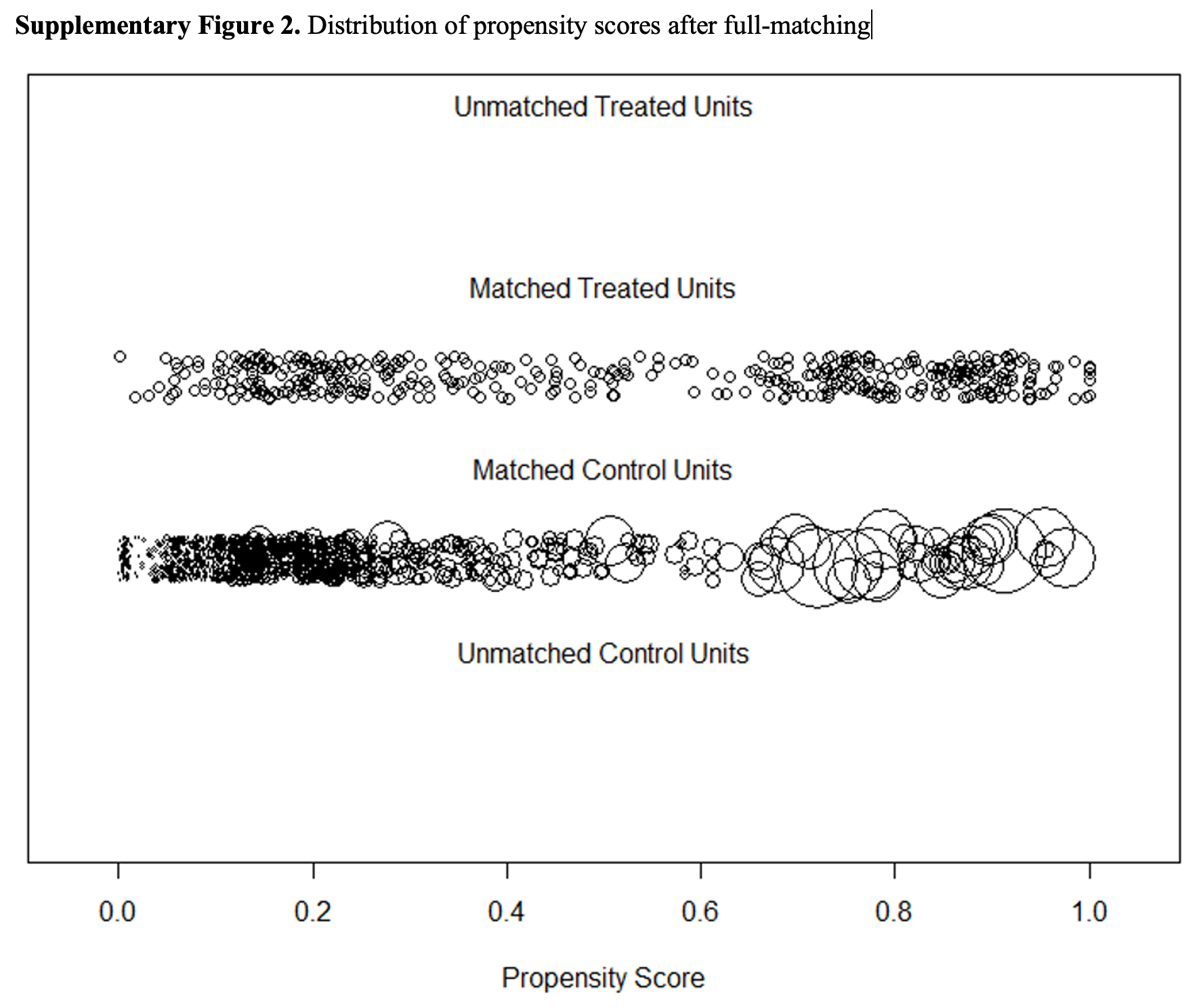
