## Supplementary file 1 for "Efficacy of Molnupiravir in reducing the risk of severe outcome in patients with SARS-CoV-2 infection: a real-life full-matched case-control study (SAVALO Study)"

### SAVALO STUDY

### TELEPHONE INTERVIEW

Dear Mr/Mrs, I am Dr … from the Unit of Infectious Diseases of the Federico II University Hospital, led by Professor Ivan Gentile. I must inform you that the present telephone call will be recorded in accordance with the privacy laws. I am calling you because it has been reported that you have had COVID-19 recently and, with your allowance, we would like to ask you some questions in order to better understand this condition. This is a telephone-only interview included in a clinical study which was granted the ethical approval by the Ethics Committee "Comitato Etico Federico II-AORN A. Cardarelli" on September 14$^{th}$ 2022.
Before you decide to give or deny your consent to the interview, I must precise that in case you decide to take part to the present study, in accordance with the European Regulation 2016/679 on the protection of natural persons with regard to the processing of personal data and on the free movement of such data and subsequent modifications, all your personal data will be pseudo-anonymised, which means that nobody outside the study group will be able to trace your identity. Moreover you will be able to withdraw your consent anytime and present a claim to the Data Protection Authority available at:.

Therefore, do you give your consent to participate to this interview?

*Option 1: Consent denied.*
Thank you very much for your time.

*Option 2: Consent granted. We offer to forward a copy of the consent by email.*

Did you receive any treatment for COVID-19 during the first week?

Yes  No

*If Yes is the answer*- Which medication did you receive?

| Medication | Dose | Duration (days) | Additional information |
|---|---|---|---|
| Azithromycin | | | |
| Dexamethasone | | | |
| Hydrocortisone | | | |
| Methylprednisolone | | | |
| Paracetamol | | | |
| Other NSAIDs (specify) | | | |
| Other medication (specify) | | | |

Did you receive any specific treatment for COVID-19?

Yes  No

*If Yes is the answer*- Which medication did you receive?

In case specific treatment for COVID-19 has been administered (Monoclonal antibodies, Paxlovid, Molnupinavir, Remdesivir) the interview must end.

*If No is the answer browse the CRF.*

*Conclusion*: Thank you very much for your time and your effort, which will help us to collect more information on COVID-19 and may prove useful to better understand how to manage this condition.

### CASE REPORT FORM

### SAVALO Study

Version 1.1

**Patient Number**:

| General Information |
|---|
| Enrollment date (DD/MM/YYYY) |
| Gender (M/F) |
| Age (years) |
| Body Weight (Kg) |

| Patient Data | |
|---|---|
| **SARS-CoV-2 Vaccination** | |
| - Completed (Yes/No) | |
| - Doses of vaccine (N°) | |
| - Last vaccination date (DD/MM/YYYY) | |
| **SARS-CoV-2 serology** | □Pos □Neg □N/A |
| - Date of the exam (DD/MM/YYYY) | |
| **Prophylaxis with Tixagevimab/Cilgavimab** | |
| - Received (Yes/No) | |
| - Doses (N°) | |
| - Last administration date (DD/MM/YYYY) | |
| **Comorbidities**<br>□ Diabetes mellitus<br>□ CKD<br>□ Haemodialysis | |

- Cardiovascular disease (specify _______________)
- COPD or other chronic lung diseases (specify _______________)
- Obesity
- Liver disease
- Neurological/Neurodegenerative disease (specify _______________)
- Autoimmune diseases (specify _______________)
- Active solid neoplasia
- Past solid neoplasia
- Active haematological malignancy
- Past
- Transplant (Specify organ transplanted __________________)
- \- Year of transplant ___________
- HIV/AIDS
- \- CD4+: □ <200 □ >200
- Other immunodeficiency (specify _______________)

| **SARS-CoV-2 infection data** | |
|---|---|
| First positive SARS-CoV-2 test (DD/MM/YYYY) | |
| Symptoms onset (DD/MM/YYYY) | |
| Reported symptoms:<br> Fever<br> Cough<br> Dyspnoea<br> Rhinitis<br> Muscle/joint pain<br> Fatigue<br> Vomit<br> Diarrhoea<br> Headache<br> Anosmia<br> Dysgeusia | |
| OSCI score* (0-8) | |
| Evidence of pneumonia at chest X-rays | □ Pos □ Neg □ N/A |

| | |
|---|---|
| □ Date of the exam (DD/MM/YYYY) | |
| Evidence of pneumonia at Chest CT scan | □ Pos (Chung score: ___) □ Neg □ N/A |
| □ Date of the exam (DD/MM/YYYY) | |
| Antiviral medications | □ None<br>□ Nirmatrelvir/r<br>□ Molnupiravir<br>□ Remdesivir short course (3 days) |
| □ Start date (DD/MM/YYYY) | |
| □ End date (DD/MM/YYYY) | |
| Other medication for COVID-19<br>□ None<br>□ Remdesivir long course (5 days)<br>□ MABS (specify _______________)<br>□ LMWH<br>□ CCS<br>□ Immunomodulators (Tocilizumab, Anakinra, Baricitinib)<br>□ Other (specify _______________) | |
| ADRs:<br>□ Fever<br>□ Dysgeusia<br>□ Nausea<br>□ Vomit<br>□ Diarrhoea<br>□ Headache<br>Other (specify ________________________________________________________________________<br>_______________________________________________________________________________)<br>ADR duration (days): ________________________________________________________ | |

| | |
|---|---|
| **Outcomes** | |
| **Day 7 evaluation** | |
| COVID-19 related symptoms still present | □ YES □ NO |
| Repeated SARS-CoV-2 test | □ POS □ NEG |

| | |
|---|---|
| OSCI Score* (0-8) | |
| Hospital admission | □ YES □ NO |
| ICU admission | □ YES □ NO |
| Death | □ YES □ NO |
| **Day 28 evaluation** | |
| COVID-19 related symptoms still present | □ YES □ NO |
| Repeated SARS-CoV-2 test | □ POS □ NEG |
| OSCI Score* (0-8) | |
| Hospital admission | □ YES □ NO |
| ICU admission | □ YES □ NO |
| Death | □ YES □ NO |
| **Additional information** | |
| *If hospital admission YES* | |
| - Admission date | |
| - Dicharge date | |
| *If hospital admission YES* | |
| - Admission to ICU date | |
| - Dicharge from ICU date | |
| *If death YES* | |
| - Date of death (DD/MM/YYYY) | |
| First negative SARS-CoV-2 test (DD/MM/YYYY) | |
| Symptoms resolution (DD/MM/YYYY) | |

***OSCI score:**

| Patient state | Description | Score |
|---|---|---|
| Uninfected | No clinical or virological evidence of infection | 0 |
| Infected but not hospitalised | Infected but no limitation of activities | 1 |
| | Limitation of activities | 2 |
| Infected and hospitalised with mild-moderate disease | Hospitalised, no oxygen therapy | 3 |
| | Oxygen therapy with mask or nasal cannula | 4 |
| Infected and hospitalised with severe disease | Non-invasive ventilation or high-flow oxygen | 5 |
| | Intubation and mechanical ventilation | 6 |
| | Ventilation and additional organ support: vasopressors, renal replacement therapy, ECMO | 7 |
| Deceased | Death | 8 |

**Legend:**

- CKD: chronic kidney disease
- COBP: chronic obstructive broncopneumopathy
- ICU: intensive care unit
- ADRs: adverse drug reactions
- MABS: monoclonal antibodies
- LMWH: low molecular weight heparin
- CCS: corticosteroids
