## Supplementary table 1 for "Efficacy of Molnupiravir in reducing the risk of severe outcome in patients with SARS-CoV-2 infection: a real-life full-matched case-control study (SAVALO Study)"

**Supplementary Table 1.** Number of patients stratified and outcome rates among included patients according to vaccination status.

| | | MNP | No MNP | p-value |
|---|---|---|---|---|
| Overall (n) | Vaccinated | 138 | 1142 | |
| | Not vaccinated | 8 | 94 | |
| *Outcomes* | | | | |
| Hospital admission (%) | Vaccinated | 3 (2.2) | 20 (1.8) | 0.730 |
| | Not vaccinated | 0 (0) | 2 (2.1) | 1.000 |
| ICU admission (%) | Vaccinated | 0 (0) | 1 (0.1) | 1.000 |
| | Not vaccinated | 0 (0) | 0 (0) | 1.000 |
| Death (%) | Vaccinated | 1 (0.7) | 15 (1.3) | 1.000 |
| | Not vaccinated | 0 (0) | 8 (8.5) | 1.000 |
| Composite outcome (%) | Vaccinated | 3 (2.2) | 31 (2.7) | 1.000 |
| | Not vaccinated | 0 (0) | 8 (8.5) | 1.000 |
